# Management of Cases Where Lower Limb Lymphoedema was Misdiagnosed as Iliac Vein Compression Syndrome

**DOI:** 10.1101/2025.04.08.25325224

**Authors:** Guijun Huo, Mingqing Du, Zhichao Yao, Dayong Zhou, Zhanao Liu

**Affiliations:** Department of Vascular surgery, The Affiliated Suzhou Hospital of Nanjing Medical University, Suzhou Municipal Hospital, Gusu School, Nanjing Medical University

**Keywords:** Iliac vein compression, secondary lymphedema, complex decongestive therapy, lymphovenous anastomosis, liposuction

## Abstract

Unilateral lower extremity lymphedema may be clinically misdiagnosed as iliac vein compression syndrome, leading to stent implantation. This study summarizes the management of cases where lower extremity lymphedema was misdiagnosed as iliac vein compression syndrome. A retrospective analysis was conducted on the edema reduction outcomes of 11 patients with secondary lymphedema who had been previously undergone iliac vein stent implantation. After these patients presented for subsequent visit and were diagnosed with lymphedema, they were treated with various therapies tailored to their specific stages of lymphedema, including complex decongestive therapy, lymphovenous anastomosis, or a combination of liposuction and lymphovenous anastomosis. Follow-up assessments of limb morphology and Lymphedema Functioning, Disability and Health Questionnaire for Lower Limb Lymphedema scores were conducted 3, 6, and 12 months post-treatment. After iliac vein stent implantation, partial relief of limb swelling was observed in six patients with stage I and IIa lymphedema, but the swelling recurred and worsened within 3 months. The remaining patients did not experience any relief of limb swelling following stent implantation. Upon revisit, four patients with Stage I and IIa lymphedema underwent complex decongestive therapy, four patients with Stage I and IIa lymphedema received lymphovenous anastomosis, and the remaining three patients with Stage IIb lymphedema underwent liposuction combined with secondary lymphovenous anastomosis. Iliac vein stent implantation does not prevent the progression of limb swelling in patients with secondary lymphedema complicated by iliac vein compression. Complex decongestive therapy, lymphovenous anastomosis, or liposuction can achieve good therapeutic outcomes for these patients.

## Introduction

The escalating incidence of malignant tumors and the enhanced survival rates of patients following radical tumor surgeries in recent years have led to a notable surge in lymphedema occurrence.^1^ Patients presenting with lower limb lymphedema at outpatient clinics may be misdiagnosed with edema resulting from other causes, such as cardiogenic, renal, or venous edema.^2,3^

Iliac vein compression syndrome is a common disease in vascular surgery. Previous studies have reported that the incidence of iliac vein compression is extremely high in the general population, reaching as high as 65%.^4,5^ The left common iliac vein often traverses the space between the aorta and lumbar vertebrae, and its compression can result in symptoms of venous outflow obstruction—a condition known as Cockett’s syndrome or May–Thurner syndrome (MTS).^6,7^ This syndrome is particularly prevalent among middle-aged women within 30–50 years of age.^8^ Patients from this age group and having postoperative lower limb lymphedema, especially in the left lower limb, due to cervical or endometrial cancer often undergo lower extremity venography when they visit vascular surgery or interventional radiology departments. Subsequently, those revealed to have iliac vein compression may be diagnosed with iliac vein compression syndrome and undergo stent implantation.

This study retrospectively reviewed 11 patients who were diagnosed with iliac vein compression syndrome and underwent stent implantation. After experiencing persistent swelling and being diagnosed with lymphedema at our hospital, these patients received specialized lymphedema treatment. The study summarizes the different treatment measures administered and their corresponding outcomes.

## Methods

### Patients

This study was performed in line with the principles of the Declaration of Helsinki. Approval was granted by the Ethics Committee of *** Hospital (IRB Number: J-2024-042-K01, dated November 25, 2024), and individual consent, including for photograph acquisition and distribution, was obtained from all included patients. The data utilized in this study were collected on 25 February 2025. The authors had access to participants’ identifiable information during the data collection phase. From January 2022 to June 2024, 165 patients with lymphedema visited the outpatient clinic or were hospitalized in *** hospital. Among them, 11 patients with lymphedema who had been previously diagnosed with iliac vein compression and undergone iliac vein stent placement were included. The patient inclusion criteria were confirmation of lymphatic obstruction through ^99^Tc^m^-DX lymphoscintigraphy, presence of unilateral secondary lymphedema, at least a 10% increase in the affected limb volume compared to that of the contralateral side, and a diagnosis of iliac vein compression and subsequent iliac vein stent implantation after lymphedema onset. The patient exclusion criteria consisted of deep venous thrombosis or iliac vein stent occlusion, inability to comply with compression therapy measures, age >80 years, recurrence of malignant tumors, and those who were lost to follow-up. The demographic information, limb circumferences, cancer surgical approach, chemical and radiation therapy, and erysipelas episodes of all included patients were recorded.

### Pre-treatment assessment

Medical history inquiry and physical examination were conducted to determine the initial diagnosis and staging of the patient’s lymphedema, which was conducted by strictly adhering to the staging system proposed by the International Society of Lymphology (ISL) **(Table 1)**.^9,10^ The morphological indicators of the patient’s limbs before iliac vein stent implantation and the effect and duration of postoperative detumescence were also investigated. Complete resolution of the affected limb to symmetry with the contralateral limb after iliac vein stent placement was defined as disease remission, whereas other situations were regarded as partial remission or ineffective treatment. All patients underwent ^99^Tc^m^-DX lymphoscintigraphy to confirm lymphatic obstruction and were screened for deep venous thrombosis or stent occlusion via vascular ultrasound or phlebography of the lower extremities. Furthermore, a limb magnetic resonance imaging scan was performed to determine whether the affected limb had fluid-predominant or solid-predominant lymphedema.^11^ Limb morphology measurements were also conducted before the patient underwent treatment to assess the severity of the condition, with all limb circumferences calculated at the midpoint, lower third, and upper third of the calves and thighs. Additionally, the frustum method was applied to estimate the lower limb volumes.^12^ Lastly, the Lymphedema Functioning, Disability and Health Questionnaire for Lower Limb Lymphedema (Lymph-ICF-LL) score and limb circumference/volume measurements were employed to establish the baseline status of the disease.^13^

**Table 1.** Staging System of the International Society of Lymphology^9,10^.

|  |  |
| --- | --- |
| 0 | Latent or subclinical lymphedema |
| I | Lymphedema that subsides with limb elevation |
| IIa | Lymphedema does not subside with limb elevation, and pitting edema is observed |
| IIb | Pitting edema is difficult to detect, while fibrosis and adiposity become prominent |
| III | Lymphostatic elephantiasis. Pitting edema is absent. Advanced stages of adiposity, fibrosis, and dermal thickening with warty overgrowths |

### Surgical Treatment

Patients with fluid-predominant stage I and IIa lymphedema were first administered complex decongestive therapy (CDT) in the lymphedema care clinic. Subsequently, patients with unsatisfactory treatment outcomes underwent peripheral lymphovenous anastomosis (LVA) after hospitalization to improve lymphatic return. LVA surgery typically involves multiple incisions, during which superficial functional lymphatic vessels are anastomosed with small veins of matching diameter under the guidance of indocyanine green lymphangiography **(Fig 1A, 1B)**. Morphological data of the limbs before and after LVA surgery were recorded **(Fig 1C, 1D)**. In the case of patients with solid-predominant stage IIb and III lymphedema, liposuction was first performed for volume reduction, followed by a second-stage LVA to enhance lymphatic return **(Fig 2A, 2B)**. Liposuction was conducted using the tumescent technique and the power-assisted liposuction device (Yangguangzhongtian, Shanxi, China) with 35 cm long cannulas. During the 3 months following the liposuction procedure, all patients wore 30–40 mmHg compression garments for limb compression. At 3 months after liposuction, LVA was performed at the inguinal region **(Fig 2C, 2D)**. Patients were provided short-stretch bandage compression for at least 3 months after LVA, followed by the application of compression garments with a pressure of 30–40 mmHg for maintenance.

**Fig 1:**
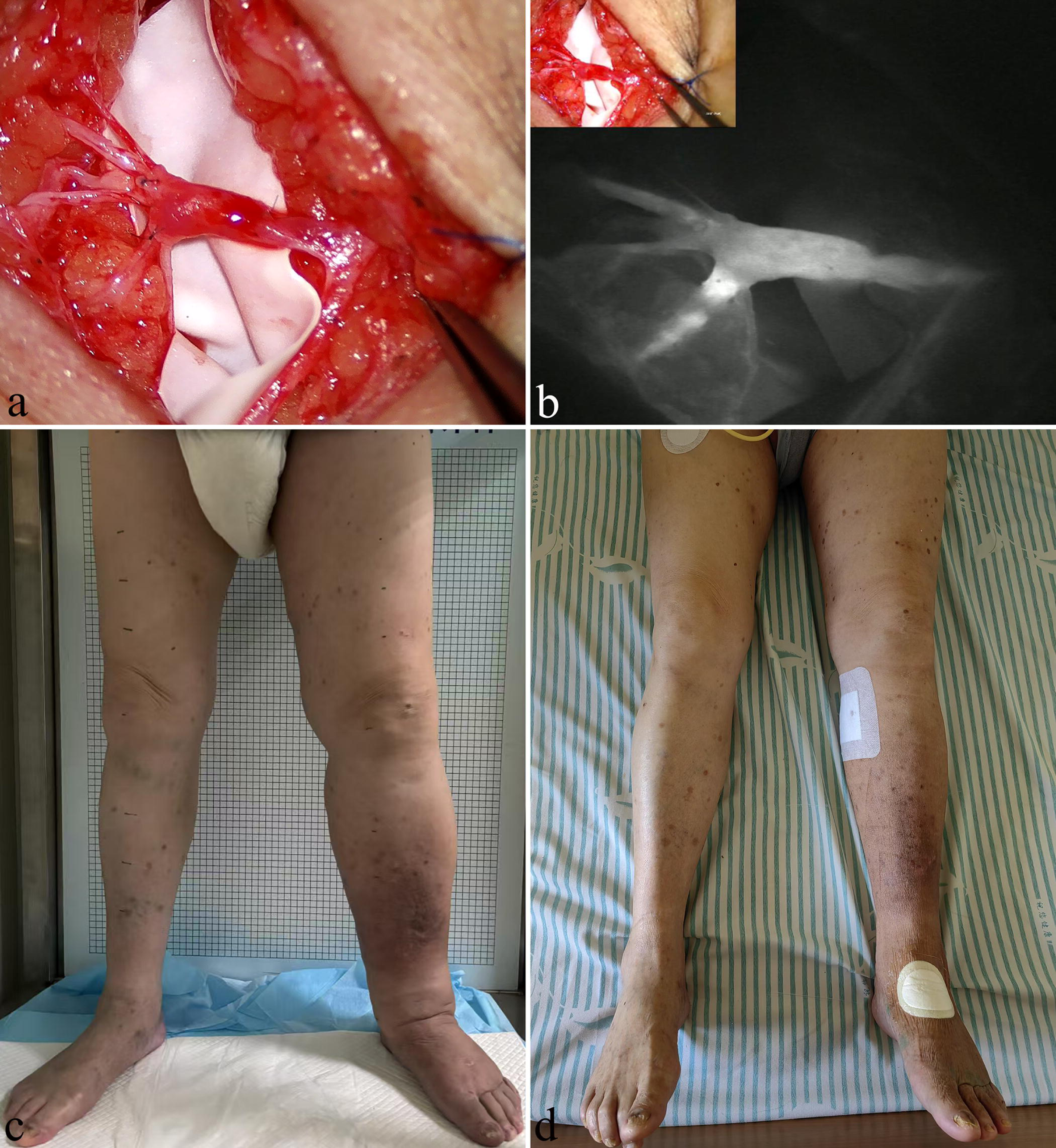
A 60s-year-old man with a history of renal tumor surgery and persistent left lower limb edema for 3 years after a prior diagnosis of left iliac vein compression syndrome and iliac vein stent implantation 1 year ago. **(A)** An incision was made on the medial side of the left calf. The valve function of the second-order branch of the great saphenous vein was found to be good, and seven lymphatic vessels were anastomosed into the vein. **(B)** Fluorescence microscopy imaging after the preoperative injection of indocyanine green lymphangiography provided clear visualization, which demonstrated smooth entry into the veins and confirmed a patent anastomosis site. **(C)** Preoperative assessment indicated that the left lower limb was swollen, with an International Society of Lymphology stage of IIa. **(D)** On the second day after surgery, the limb swelling significantly improved without complex decongestive therapy.

**Fig 2:**
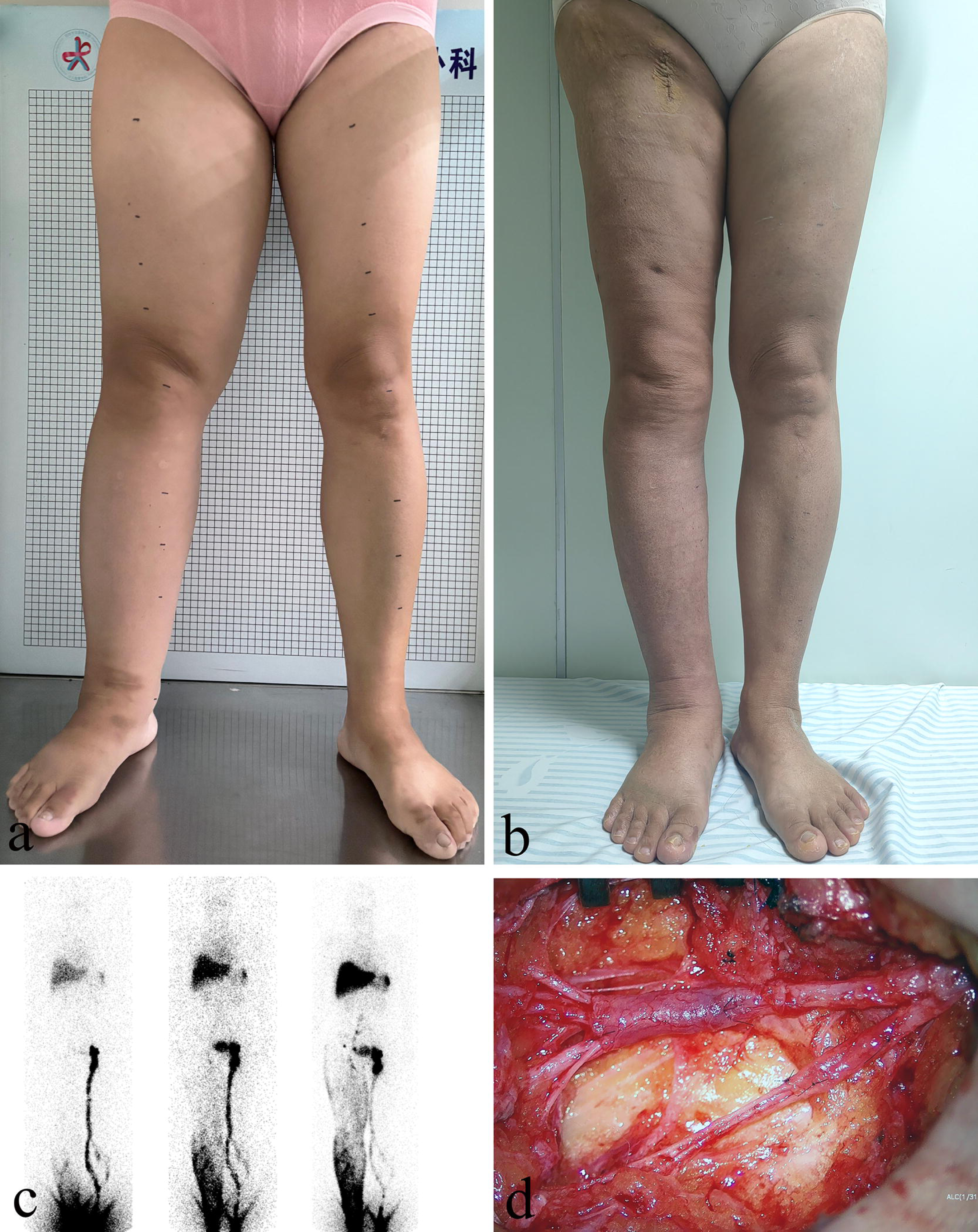
A 50s-year-old woman with a history of cervical cancer surgery was diagnosed with right iliac vein compression 4 years ago and underwent stent implantation with no swelling relief. **(A)** Prior to the surgical procedure during the hospitalization stage, the patient’s right lower limb showed significant swelling that was classified as International Society of Lymphology stage IIb. **(B)** After liposuction followed by lymphovenous anastomosis 3 months later, the swelling in the patient’s limb significantly improved. **(C)** Preoperative ^99^Tc^m^-DX lymphoscintigraphy in the anterior position at 10 min, 1 h, and 3 h post-injection demonstrated lymphatic return obstruction in the right lower limb. **(D)** The lymphatic vessels, lateral femoral vein, and its branches were dissected at the inguinal level. After confirming good valve function, the lymphatic vessels were anastomosed to the lateral femoral vein and its branches.

### Follow up

During discharge, all complications were comprehensively recorded for each patient and categorized into major complications requiring surgical intervention and minor complications that could be alleviated through conservative treatment. All postoperative patients and those receiving compression therapy in the outpatient setting underwent follow-up assessments at 3, 6, and 12 months. During these assessments, the patients received guidance on compression therapy, underwent Lymph-ICF-LL scoring, and had their limb morphology measurements recorded. The volume difference between bilateral limbs and the difference rate after the treatment were also compared. The volume difference rate of the bilateral limbs before and after lymphedema treatment was defined as follows: (volume of the affected limb − volume of the healthy limb)/volume of the healthy limb.

### Statistical analysis

Statistical analysis was performed using SPSS statistical software (version 17; SPSS, Chicago, Illinois). Kolmogorow-Smironov test was used to confirm data normality. Quantitative variables were summarized as mean and standard deviation (SD) if normally distributed or median and interquartile range (IQR). Categorical variables were presented as numbers and percentages. Pre and post-treatment Lymph-ICF-LL was analyzed with paired t-tests. A value of *P* ≤0.05 was considered to be significant.

## Results

A total of 11 patients were included in this study, with an average age of 63.18 ± 9.89 years and consisting of nine women and two men. The baseline characteristics of the patients are presented in **Table 2**. Five patients had received treatment with compression stockings, whereas none had undergone systematic CDT before hospitalization. Moreover, none of the patients exhibited signs of superficial varicose veins, lipodermatosclerosis, or ulcers. Ten patients underwent iliac vein stent implantation in an external hospital, while one received the procedure in the vascular surgery department of our hospital **(Fig 3A, 3B)**. Specifically, nine patients were treated with left iliac vein stent implantation due to left iliac vein stenosis, and two underwent stent implantation for right iliac vein stenosis. Six patients with stage I and IIa lymphedema experienced partial relief of limb swelling following iliac vein stent implantation; however, the swelling recurred and continued to progress within 3 months. Additionally, no relief of limb swelling was observed in the remaining five patients after stent implantation. The clinical data of all 11 patients are provided in **Table 3**.

**Fig 3:**
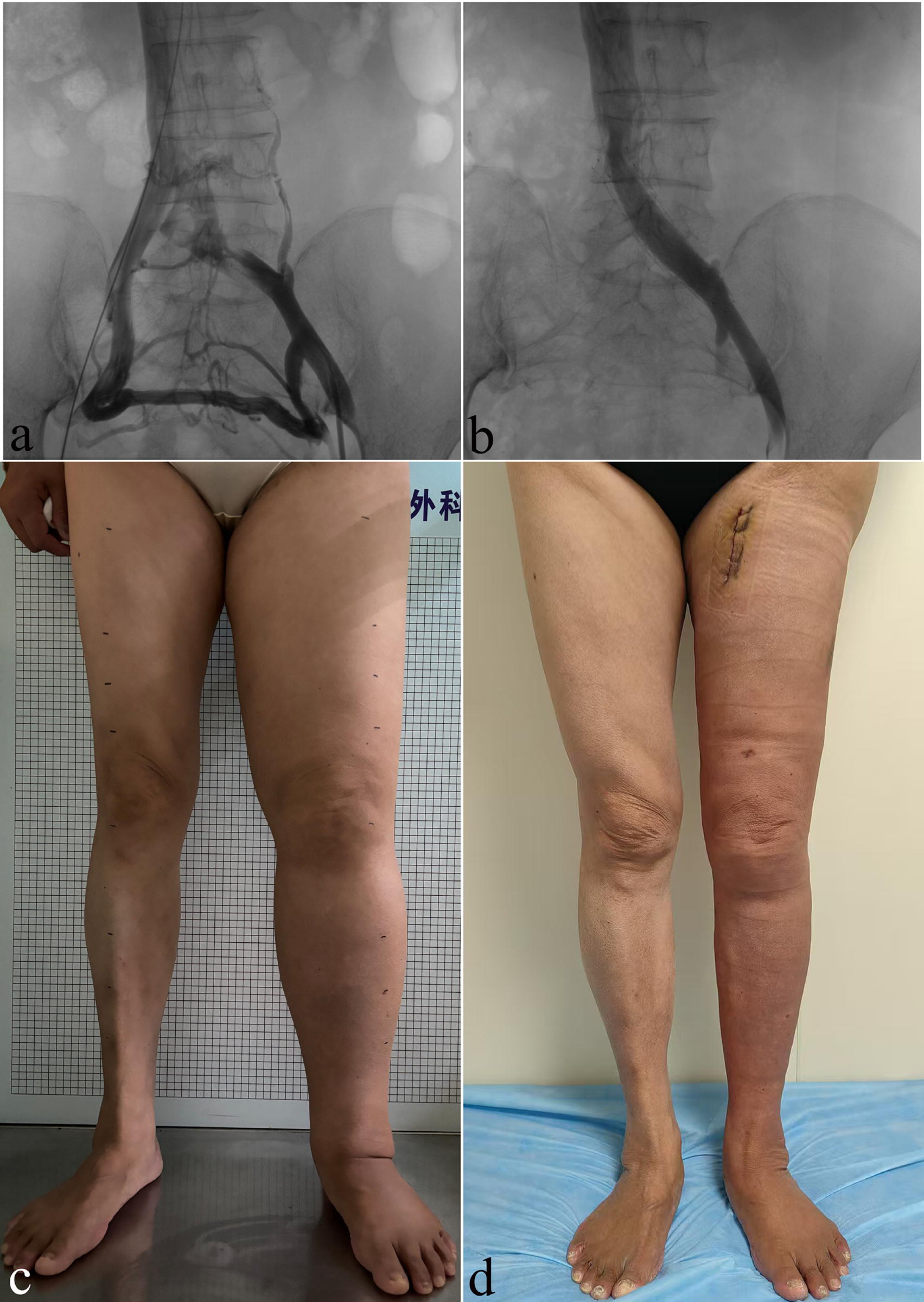
A 40s-year-old woman who had a history of cervical cancer surgery was diagnosed with left iliac vein compression syndrome 18 months earlier and underwent stent implantation. However, the swelling symptoms did not show any improvement. **(A)** Left iliac venography revealed severe stenosis of the left common iliac vein accompanied by extensive collateral vessel formation, with blood flow compensation via the contralateral internal iliac vein. **(B)** Follow-up venography revealed smooth blood flow in the left common iliac vein after stent implantation and the disappearance of the collateral veins. **(C)** Before the surgical procedure during the hospitalization stage, the patient’s left lower limb demonstrated significant swelling that was categorized as International Society of Lymphology stage IIb. **(D)** This female patient experienced immediate reduction in the volume of her swollen limb after undergoing liposuction.

**Table 2.** Demographics and baseline characteristics of the study patients.

| Patient number | Gender | BMI (kg/m<sup>2</sup>) | Affected limb | Primary malignancy | Radiation therapy | Lymphedema duration (months) | Cellulitis history |
| --- | --- | --- | --- | --- | --- | --- | --- |
| 1 | Female | 24 | Left | Cervical cancer | Not performed | 45 | Single episode |
| 2 | Female | 25.5 | Left | Cervical cancer | Not performed | 60 | Never |
| 3 | Female | 25.2 | Right | Cervical cancer | Performed | 60 | Recurrent episode |
| 4 | Male | 24.5 | Left | Renal cell cancer | Not performed | 36 | Recurrent episode |
| 5 | Female | 24.4 | Left | Endometrial cancer | Not performed | 12 | Never |
| 6 | Female | 18.7 | Left | Endometrial cancer | Not performed | 10 | Single episode |
| 7 | Female | 24.7 | Left | Cervical cancer | Not performed | 24 | Never |
| 8 | Female | 24.6 | Right | Cervical cancer | Performed | 120 | Recurrent episode |
| 9 | Female | 29.3 | Left | Rectal cancer | Not performed | 6 | Never |
| 10 | Female | 21.1 | Left | Cervical cancer | Not performed | 14 | Single episode |
| 11 | Male | 24.2 | Left | Rectal cancer | Performed | 36 | Never |

BMI, body mass index.

**Table 3.**
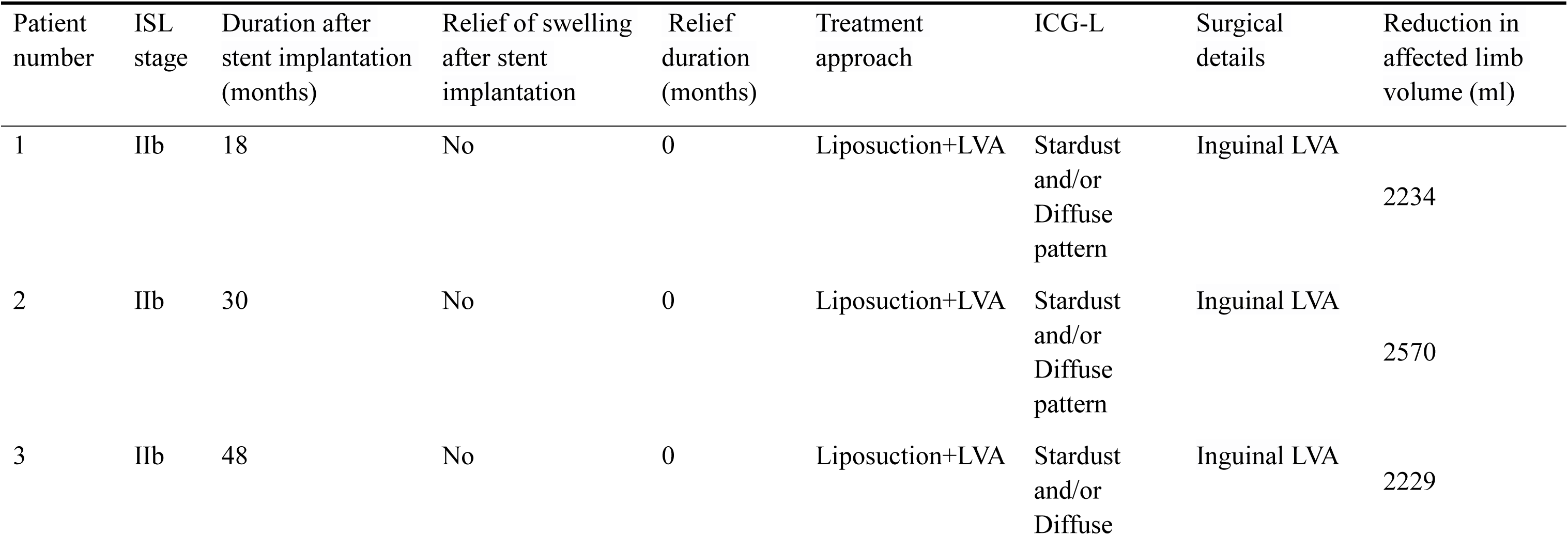
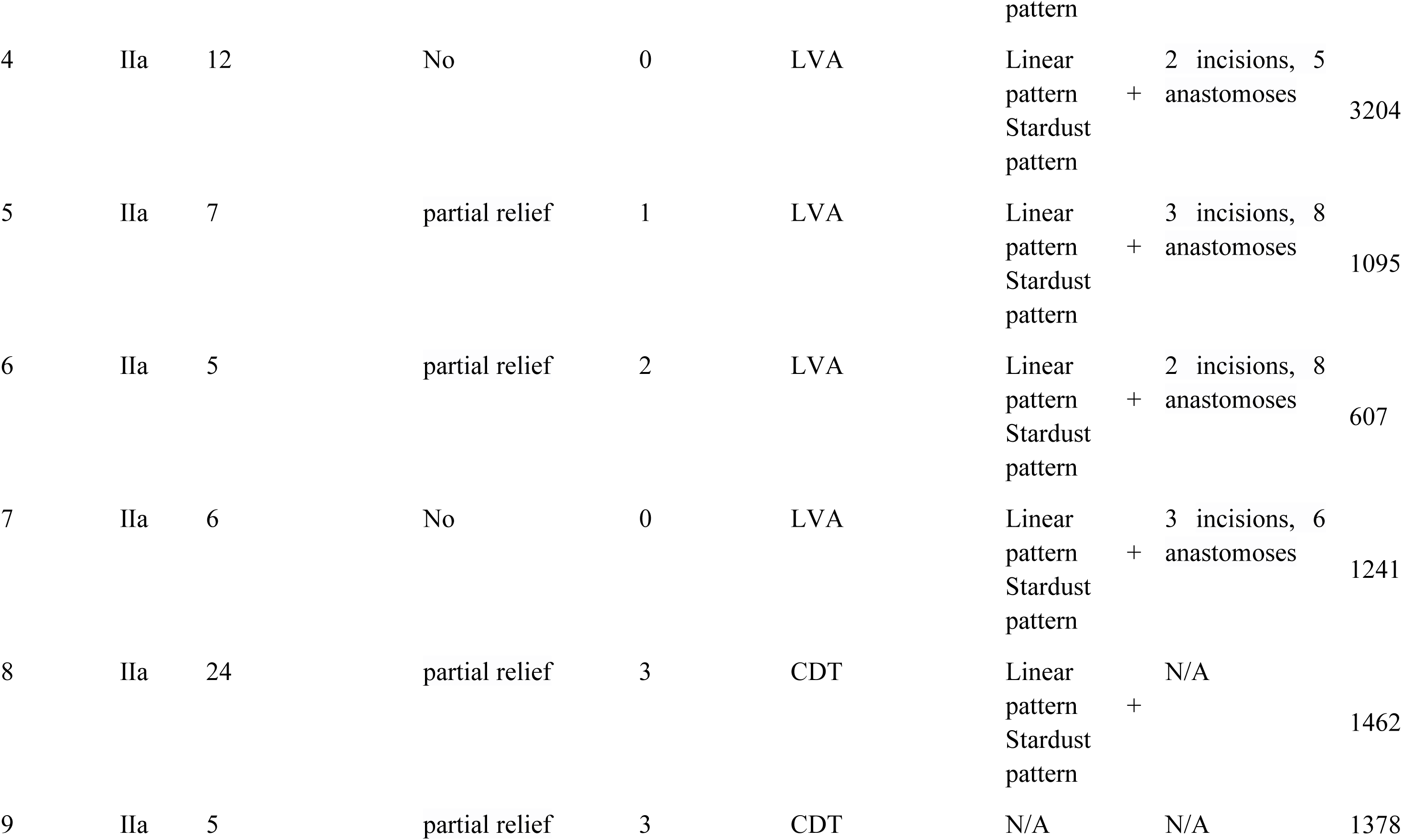
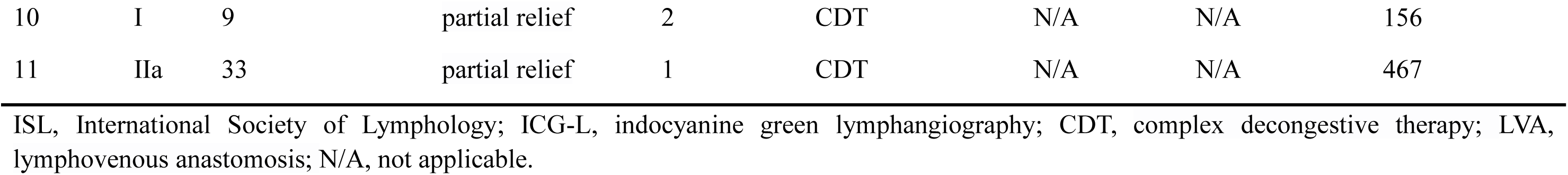
Clinical data of the study patients with secondary lymphedema complicated by iliac vein compression.

All included patients were confirmed to have patent iliac vein stents via ultrasonography or angiography and were diagnosed with lymphedema through ^99^Tc^m^-DX lymphoscintigraphy. Four patients with stage I and IIa lymphedema achieved satisfactory outcomes with CDT, thereby not requiring further surgical intervention. These four patients showed a median volume reduction of 923.26 (234.11, 1441.28) ml and a decrease in the median volume difference (volume difference rate) from 1469.16 mL (22%) to 545.90 mL (8%), along with a decline in the median Lymph-ICF-LL score from 155 to 56 that indicated an improved quality of life.

Four other patients with stage I and IIa lymphedema who responded poorly to CDT underwent LVA during their hospitalization, with no major or minor complications observed postoperatively. These patients experienced a median volume reduction of 1168.72 (729.53, 2714.14) ml over a follow-up period of 6 months and a decrease in the median volume difference (volume difference rate) from 1904.30 mL (30%) to 735.58 mL (12%), while their median Lymph-ICF-LL score decreased from 168 to 66.

The remaining three patients with stage IIb lymphedema were admitted for surgical intervention after unsatisfactory results from CDT. The surgical approach involved initial liposuctionl **(Fig 3C, 3D)**, followed by a second-stage LVA after 3 months to improve lymphatic drainage. During the 6-month follow-up period, these patients presented with a median volume reduction of 2234.31 ml, a decrease in the median volume difference (volume difference rate) from 3013.82 mL (43%) to 784.39 mL (11%), and a decline in the median Lymph-ICF-LL score from 158 to 59. Follow-up data of all included patients are presented in **Table 4**.

**Table 4.** Clinical data of the bilateral limbs of the study patients before and after lymphedema treatment.

| CDT (n = 4) |  |  |  | LVA (n = 4) |  |  | Liposuction+LVA (n = 3) |  |  |
| --- | --- | --- | --- | --- | --- | --- | --- | --- | --- |
| Volume difference between the bilateral limbs before and after lymphedema treatment (ml) |  |  |  |  |  |  |  |  |  |
| Time point | Min | Max | M (P<sub>25</sub>,P<sub>75</sub>) | Min | Max | M (P<sub>25</sub>,P<sub>75</sub>) | Min | Max | M (P<sub>25</sub>,P<sub>75</sub>) |
| Before treatment | 280.14 | 2711.19 | 1469.16(367.50, 2610.58) | 717.98 | 4206.48 | 1904.30(964.51, 3681.00) | 2358.86 | 3425.01 | 3013.82(2358.86, -) |
| 6 months after treatment | 123.95 | 1249.03 | 545.90(133.39, 1169.30) | 110.45 | 1001.60 | 735.58(234.98, 966.85) | 124.55 | 854.76 | 784.39(124.55, -) |
| Volume difference rate of the bilateral limbs before and after lymphedema treatment |  |  |  |  |  |  |  |  |  |
| Time point | Min | Max | M (P<sub>25</sub>,P<sub>75</sub>) | Min | Max | M (P<sub>25</sub>,P<sub>75</sub>) | Min | Max | M (P<sub>25</sub>,P<sub>75</sub>) |
| Before treatment | 0.04 | 0.44 | 0.22 (0.05, 0.42) | 0.17 | 0.58 | 0.30 (0.20, 0.51) | 0.39 | 0.68 | 0.43 (0.39, -) |
| 6 months after treatment | 0.02 | 0.20 | 0.08 (0.02, 0.19) | 0.03 | 0.14 | 0.12 (0.05, 0.14) | 0.02 | 0.17 | 0.11 (0.02, -) |
| Lymph-ICF-LL score before and after lymphedema treatment (points) |  |  |  |  |  |  |  |  |  |
| Time point | Min | Max | M (P<sub>25</sub>,P<sub>75</sub>) | Min | Max | M (P<sub>25</sub>,P<sub>75</sub>) | Min | Max | M (P<sub>25</sub>,P<sub>75</sub>) |
| Before treatment | 150 | 161 | 155 (150, 160) | 142 | 200 | 168 (146, 195) | 153 | 189 | 158 (153, -) |
| 6 months after treatment | 42 | 73 | 56 (45, 69) | 39 | 76 | 66 (44, 76) | 43 | 62 | 59 (43, -) |

CDT, complex decongestive therapy; LVA, lymphovenous anastomosis; Lymph-ICF-LL, Lymphedema Functioning, Disability and Health
Questionnaire for Lower Limb Lymphedema.

All patients with lymphedema who received CDT treatment and surgical intervention showed reduced limb volume, particularly after liposuction. However, one patient experienced a postoperative complication of localized skin necrosis (approximately 4 cm^2^) after the liposuction procedure, while others did not develop complications such as lymphatic leakage, wound dehiscence, or infection. During the follow-up period, all patients showed an improvement in their quality of life. Lastly, all patients wore compression garments with a pressure of 30–40 mmHg to maintain the therapeutic effect.

## Discussion

The primary cause of secondary lymphedema in the lower limbs is attributed to lymph node dissection and radiation therapy associated with uterine cancer, ovarian cancer, and other malignant tumors. Previous studies have reported that approximately 50% of patients develop lower extremity lymphedema ≥5 years after receiving gynecologic cancer treatment.^14^ The long-term accumulation of nutrient-rich lymph in the interstitial spaces can induce tissue edema. Further progression can promote fat deposition, fibrosis, and recurrent erysipelas, ultimately affecting limb appearance, causing malformation and dysfunction, and severely affecting the patients’ quality of life and mental health.^15,16^ Moreover, the proliferation of skin and subcutaneous tissues generates more lymph, which creates a vicious cycle that gradually worsens the disease condition.^17^

The ISL has identified and described four clinical stages of lymphedema.^9,18^ According to the different pathological stages, surgical treatment for lymphedema primarily involves debulking procedures and lymphatic reconstruction surgeries. Particularly, fluid-predominant stage I and IIa lymphedema can be treated using surgical techniques that enhance lymphatic drainage. These methods include LVA, vascularized lymph node transfer/transplantation, and lymphatic grafting. In solid-predominant stage IIb and III lymphedema, a combination of debulking procedures and lymphatic reconstruction surgeries is recommended.^11,19^ However, centers capable of comprehensively performing surgical treatments for lymphedema are limited, thus making it challenging for patients to receive appropriate and effective surgical interventions. Occasionally, patients with stage I or IIa lymphedema of the lower extremities with a primary manifestation of pitting edema may be misdiagnosed with venous edema after consulting vascular surgeons. Further imaging examinations of these patients may indicate a diagnosis of iliac vein compression syndrome, followed by treatment with iliac venoplasty.

Prior cadaveric and radiographic studies have reported that the prevalence of iliac vein compression is remarkably high in the general population.^4,5^ Additionally, MTS is predominant among women between the ages of 30 and 50, whereas symptomatic cases may present with venous insufficiency symptoms such as limb swelling, varicose veins, and even venous ulcers.^8^ The age of postoperative lymphedema onset in gynecological malignant tumors is also approximately 50 years of age, which highlights the need to distinguish it from MTS characterized by limb swelling. Occasionally, patients with unilateral limb swelling who are treated using vascular surgery undergo a preliminary examination via lower limb vascular ultrasound, followed by anterograde lower limb venous angiography or computed tomography venography (CTV) to identify deep vein problems. After finding obvious iliac vein stenosis accompanied by obvious pelvic vein collateral on anterograde angiography or CTV of the lower limb vein and considering the swelling of the patient’s limbs, angioplasty and stenting are performed to alleviate the limb swelling.

In this study, six out of eight patients with stage I and IIa lymphedema complicated by iliac vein compression syndrome who underwent iliac vein stent implantation experienced varying degrees of edema relief. Although the patients gained certain short-term benefits from the surgery, the progressive course of lymphedema was not altered. Moreover, the remaining three patients with stage IIb lymphedema did not experience relief from limb swelling after iliac vein stenting. Therefore, patients who present with lower extremity edema and imaging confirming iliac vein compression should be carefully questioned for a history of radical surgery for gynecological malignancies or other surgeries. Subsequently, a differential diagnosis of lymphedema should be considered in patients with relevant medical history, and stent implantation should be approached with greater caution. In cases where iliac vein compression is detected through imaging examinations, the decision to perform stent implantation depends on the degree of stenosis, clinical symptoms, and Clinical, Etiology, Anatomy, and Pathophysiology (CEAP) classification.^20^ According to the guidelines proposed by the American Venous Forum, Society for Vascular Surgery, European Society for Vascular Surgery, and Society of Interventional Radiology, stent implantation is recommended for patients who have clinically relevant venous outflow obstruction classified as CEAP 3–6, along with venous claudication, pelvic pain, and morphological indication of >50% area reduction.^21–23^ Specifically, C3 refers to edema, C4 to pigmentation or lipodermatosclerosis, C5 to healed venous ulcer, and C6 to active venous ulcer. Iliac vein stenting is the appropriate first-line treatment for symptomatic patients with obstructive disease, specifically those with skin or subcutaneous changes and healed or active ulcers (i.e., CEAP 4–6). Stenting may also be considered for patients with edema due to venous disease (i.e., CEAP 3), provided careful clinical judgment is exercised because of the potential for varied coexisting nonvenous causes of edema.

However, the recommendations in the previously mentioned guidelines are all based on venous-related diseases. The detection of iliac vein stenosis on imaging examinations in patients with lymphedema may indicate venous-lymphatic mixed edema rather than purely venous-related edema. Therefore, stent implantation should be considered with even greater caution in such patients. However, patients with stage I and II lymphedema and concurrent significant iliac vein compression on venography examination may still achieve some benefits through iliac vein stent implantation. Thus, the rationality of this procedure cannot be completely negated. Relief of venous obstruction can also improve lymphatic stagnation to some extent. Moreover, subsequent LVA requires a relatively normal venous return status and cannot be performed under the conditions of an obstructed venous outflow tract. Furthermore, veins with good valve function should be selected to avoid venous reflux, which can cause thrombosis at the anastomotic site between the lymphatic ducts and veins.^17,24,25^ In the present study, patients with lymphedema were initially treated using conservative therapy, specifically systematic CDT. Conservative therapies, such as the application of short-stretch bandages, compression garments, and manual lymphatic drainage, constitute the first-line treatment for lymphedema of the extremities.^9^ Most patients, particularly those with early-stage lymphedema, can achieve satisfactory results with CDT. LVA serves as an effective treatment option in patients with stage I or IIa lymphedema who have failed conservative treatment, experienced recurrent lymphangitis, or are highly dependent on compressive therapy measures. LVA is a microsurgical technique that creates a bypass between submillimeter-sized lymphatic channels and veins to improve lymphatic drainage, reduce edema volume, and alleviate related symptoms.^19,26^ LVA is a safe and effective treatment method, which is minimally invasive and does not cause severe complications.^27,28^ In the case of patients with stage IIb or III lymphedema characterized by substantial fat deposition, liposuction is employed to rapidly reduce limb volume, followed by inguinal LVA in the second stage to augment lymphatic flow. The satisfactory treatment outcomes observed in the patients with lymphedema in this study can be attributed to the appropriate surgical options implemented based on their distinct pathophysiological changes.^27,28^

However, this study has certain limitations that should be considered. First, most patients in this study underwent iliac vein stenting in other hospitals, which resulted in the lack of precise assessments of preoperative and postoperative limb morphology.

Second, the number of patients in this study was relatively small, which restricted our investigation to merely performing an efficacy analysis of iliac vein stenting and specialized lymphedema treatment without conducting subgroup analyses to compare the different treatment approaches.

## Conclusion

Iliac vein stent implantation does not prevent lymphedema progression in patients with secondary lymphedema complicated by iliac vein compression. Although iliac vein stent implantation can create better venous conditions for subsequent LVA, the indications for iliac vein stenting should be more strictly controlled. LVA or liposuction based on the different pathophysiological stages of secondary lymphedema can achieve favorable therapeutic outcomes.

## Data Availability

The datasets analysed during the current study are available from the corresponding author on reasonable request.

## Acknowledgements

None.

## References

1. Will P, Dragu A, Zuther J, Heil J, Chang DH, Traber J, et al. Evidenz der modernen Diagnostik, der konservativen und chirurgischen Therapie des sekundären Lymphödems [Evidence of modern diagnostic, conservative, and surgical therapy of secondary lymphoedema]. Handchir Mikrochir Plast Chir. 2024 Aug;56(4):291–300.

2. Wollina U, Heinig B. Differenzialdiagnostik von Lipödem und Lymphödem : Ein Leitfaden für die Praxis [Differential diagnostics of lipedema and lymphedema : A practical guideline]. Hautarzt. 2018;69(12):1039–1047.

3. Vignes S. Les lymphœdèmes : du diagnostic au traitement [Lymphedema: From diagnosis to treatment]. Rev Med Interne. 2017;38(2):97–105.

4. Zucker EJ, Ganguli S, Ghoshhajra BB, Gupta R, Prabhakar AM. Imaging of venous compression syndromes. Cardiovasc Diagn Ther. 2016;6(6):519–532.

5. Harbin MM, Lutsey PL. May-Thurner syndrome: History of understanding and need for defining population prevalence. J Thromb Haemost. 2020;18(3):534–542.

6. Poyyamoli S, Mehta P, Cherian M, Anand RR, Patil SB, Kalva S, et al. May-Thurner syndrome. Cardiovasc Diagn Ther. 2021 Oct;11(5):1104–1111.

7. Mangla A, Hamad H. May-Thurner Syndrome. In: StatPearls. Treasure Island (FL): StatPearls Publishing; March 11, 2024.

8. MAY R, THURNER J. The cause of the predominantly sinistral occurrence of thrombosis of the pelvic veins. Angiology. 1957;8(5):419–427.

9. Executive Committee of the International Society of Lymphology. The diagnosis and treatment of peripheral lymphedema: 2020 Consensus Document of the International Society of Lymphology. Lymphology. 2020;53(1):3–19.

10. Doubblestein D, Campione E, Hunley J, Schaverien M. Pre- and Post-Microsurgical Rehabilitation Interventions and Outcomes on Breast Cancer-Related Lymphedema: a Systematic Review. Curr Oncol Rep. 2023;25(9):1031–1046.

11. Knackstedt R, Chen WF. Current Concepts in Surgical Management of Lymphedema. Phys Med Rehabil Clin N Am. 2022;33(4):885–899.

12. Sitzia J. Volume measurement in lymphoedema treatment: examination of formulae. Eur J Cancer Care (Engl). 1995;4(1):11–16.

13. Devoogdt N, De Groef A, Hendrickx A, Damstra R, Christiaansen A, Geraerts I, et al. Lymphoedema Functioning, Disability and Health Questionnaire for Lower Limb Lymphoedema (Lymph-ICF-LL): reliability and validity. Phys Ther. 2014 May;94(5):705–21.

14. Kuroda K, Yamamoto Y, Yanagisawa M, Kawata A, Akiba N, Suzuki K, et al. Risk factors and a prediction model for lower limb lymphedema following lymphadenectomy in gynecologic cancer: a hospital-based retrospective cohort study. BMC Womens Health. 2017;17(1):50.

15. Tashiro K, Feng J, Wu SH, Mashiko T, Kanayama K, Narushima M, et al. Pathological changes of adipose tissue in secondary lymphoedema. Br J Dermatol. 2017;177(1):158–167.

16. Ridner SH. Pathophysiology of lymphedema. Semin Oncol Nurs. 2013;29(1):4–11.

17. Chang K, Xia S, Liang C, Sun Y, Xin J, Shen W. A clinical study of liposuction followed by lymphovenous anastomosis for treatment of breast cancer-related lymphedema. Front Surg. 2023;10:1065733.

18. Executive Committee. The Diagnosis and Treatment of Peripheral Lymphedema: 2016 Consensus Document of the International Society of Lymphology. Lymphology. 2016;49(4):170–184.

19. Gallagher K, Marulanda K, Gray S. Surgical Intervention for Lymphedema. Surg Oncol Clin N Am. 2018;27(1):195–215.

20. Lurie F, Passman M, Meisner M, Dalsing M, Masuda E, Welch H, et al. The 2020 update of the CEAP classification system and reporting standards. J Vasc Surg Venous Lymphat Disord. 2020;8(3):342–352.

21. De Maeseneer MG, Kakkos SK, Aherne T, Baekgaard N, Black S, Blomgren L, et al. Editor’s Choice - European Society for Vascular Surgery (ESVS) 2022 Clinical Practice Guidelines on the Management of Chronic Venous Disease of the Lower Limbs. Eur J Vasc Endovasc Surg. 2022 Feb;63(2):184–267.

22. Masuda E, Ozsvath K, Vossler J, Woo K, Kistner R, Lurie F, et al. The 2020 appropriate use criteria for chronic lower extremity venous disease of the American Venous Forum, the Society for Vascular Surgery, the American Vein and Lymphatic Society, and the Society of Interventional Radiology. J Vasc Surg Venous Lymphat Disord. 2020;8(4):505–525.e4.

23. Villalba LM, Bayat I, Dubenec S, Puckridge P, Thomas S, Varcoe R, et al. Review of the literature supporting international clinical practice guidelines on iliac venous stenting and their applicability to Australia and New Zealand practice. J Vasc Surg Venous Lymphat Disord. 2024;12(5):101843.

24. Ito R, Wu CT, Lin MC, Cheng MH. Successful treatment of early-stage lower extremity lymphedema with side-to-end lymphovenous anastomosis with indocyanine green lymphography assisted. Microsurgery. 2016;36(4):310–315.

25. Campisi C, Bellini C, Campisi C, Accogli S, Bonioli E, Boccardo F. Microsurgery for lymphedema: clinical research and long-term results. Microsurgery. 2010;30(4):256–260.

26. Yoo MY, Woo KJ, Kang SY, Moon BS, Kim BS, Yoon HJ. Efficacy of preoperative lymphoscintigraphy in predicting surgical outcomes of lymphaticovenous anastomosis in lower extremity lymphedema: Clinical correlations in gynecological cancer-related lymphedema. PLoS One. 2024;19(1):e0296466.

27. Hamada E, Onoda S, Satake T. Efficacy of Lymphaticovenular Anastomosis for Secondary Upper Extremity Lymphedema: Treatment Strategies with Effects of Compression Therapy Discontinuation and Site-Specific Evaluation of the Upper Extremity. Lymphat Res Biol. 2023;21(6):574–580.

28. Koshima I, Nanba Y, Tsutsui T, Takahashi Y, Itoh S. Long-term follow-up after lymphaticovenular anastomosis for lymphedema in the leg. J Reconstr Microsurg. 2003;19(4):209–215.

